# Functional outcome, return to work and quality of life in patients with non-aneurysmal subarachnoid hemorrhage

**DOI:** 10.1101/2024.12.18.24319292

**Authors:** Wouter J. Dronkers, Menno R. Germans, René Post, Bert A. Coert, Jonathan M. Coutinho, René van den Berg, W. Peter Vandertop, Dagmar Verbaan

**Affiliations:** Amsterdam UMC location University of Amsterdam, department of Neurosurgery, Meibergdreef 9, Amsterdam, the Netherlands; Amsterdam Neuroscience Centre, Neurovascular Disease, Meibergdreef 9, Amsterdam, the Netherlands; University Hospital Zurich, University of Zurich, department of Neurosurgery, Zurich, Switzerland; Clinical Neuroscience Centre, Zurich, Switzerland; Amsterdam UMC location University of Amsterdam, department of Neurology, Meibergdreef 9, Amsterdam, the Netherlands; Amsterdam UMC location University of Amsterdam, department of Radiology and Nuclear Medicine, Meibergdreef 9, Amsterdam, the Netherlands

**Author notes:** **Corresponding author:** Wouter J. Dronkers, MD LLM MA, Department of Neurosurgery, Amsterdam UMC Location University of Amsterdam, Meibergdreef 9, room H2-259, 1105 AZ Amsterdam, The Netherlands, **Email:**.

**Keywords:** complications, neurology, stroke, subarachnoid hemorrhage, outcome, return to work

## Abstract

**Background:** Non-aneurysmal, non-traumatic subarachnoid hemorrhage (nSAH) refers to cases where a causative aneurysm cannot be identified. We studied six-months’ outcomes in nSAH patients.

**Methods:** From a prospective SAH registry of all nSAH patients admitted between 2012 and 2023, relevant complications and outcomes were collected. Functional outcome and return-to-work at six months were assessed using the modified Rankin Scale (mRS), quality of life with the EuroQol-5Dimensions (EQ-5D) and Hospital Anxiety & Depression Scale (HADS), and an institutional 14-item questionnaire for assessment of residual symptoms.

**Results:** 325 consecutive nSAH patients were included (191 non-perimesencephalic, non-aneurysmal subarachnoid hemorrhage [NPSAH]; 134 perimesencephalic subarachnoid hemorrhage [PMSAH]). 303 (93%; 179 NPSAH and 124 PMSAH) were available at follow-up (7 patients died). Favorable functional outcome (mRS-score 0-2) was reported in 271 (89%) patients and did not differ between NPSAH- and PMSAH. 141 (77%) patients returned to work, whereas only 71 (39%) patients reached their previous level of work. PMSAH patients were more likely to return to work (68/96 (71%) NPSAH and 73/87 (84%) PMSAH, respectively, p=0.01). Furthermore, PMSAH patients were more likely to fully return to work (p=.034). The mean (SD) EQ-5D and EQ-VAS scores were 0.827 (0.184) and 74 (16), respectively. The HADS-A and -D scores were deviant (score >7 points) in 53 (23%) and 48 (21%) patients, respectively. Only 39 patients (16%) denied experiencing residual symptoms. Increased fatigue (n=164; 68%), increased concentration difficulties (n=130; 54%), and increased forgetfulness (n=121; 50%) were the most frequently reported residual symptoms.

**Conclusions:** This study reveals that the majority of nSAH patients reports residual symptoms and did not return to their previous level of work at six months follow-up, despite a favorable functional outcome. These findings nuance the perception of a good outcome, as suggested in previous studies, warranting further research on possible rehabilitative interventions and counseling in these patients.

## Introduction

Non-aneurysmal subarachnoid hemorrhage (nSAH) accounts for 15% of all non-traumatic subarachnoid hemorrhages ^1,2^. In contrast to aneurysmal SAH (aSAH), nSAH is characterized by a more favorable clinical course and a better prognosis at three- and six-months follow-up ^3^. Among nSAH patients, perimesencephalic subarachnoid hemorrhage (PMSAH) tends to have a better functional outcome than non-perimesencephalic subarachnoid hemorrhage (NPSAH) ^4–6^. This difference may be attributed to the substantial variation in hemorrhage volume and, more notably, the occurrence of hydrocephalus, delayed cerebral ischemia, other complications associated with hospitalization, and case fatality ^4,7,8^.

The assessment of outcomes in patients following a subarachnoid hemorrhage (SAH) uses established scales such as the modified Rankin Scale (mRS), Glasgow Outcome Scale (GOS) or the GOS extended scale (GOSE) ^9^. Favorable outcome is commonly defined as an mRS-score 0-2, a GOS-score 4-5, or a GOSE-score 5-8 ^10^. In nSAH patients, the determination of a favorable outcome is often contextualized by a comparison to aSAH ^3,11^. However, these scales frequently place considerable emphasis on physical disability and dependency, whilst these may not manifest to the same extent in nSAH patients as observed in those with true aneurysmal SAH. Other sequelae, such as the ability to return to work, symptoms of anxiety and depression, and overall decreased quality of life, may also be prevalent and, if overlooked, result in an underestimation, and subsequent under-treatment, of patients, particularly after hospital discharge ^12^.

Prospective studies in the nSAH population have mostly focused on functional outcome ^8,13,14^. Studies reporting on residual symptoms, return-to-work after the hemorrhage and quality of life, are relatively sparse, leaving these domains underreported ^15–17^. Moreover, previous studies included relatively small sample sizes, or were conducted in a retrospective fashion with variable, and often short, follow-up times ^18–23^. The aim of the present study is to determine the six months outcome in a large prospective cohort of patients after a non-aneurysmal, non-traumatic subarachnoid hemorrhage with a focus on physical disability, return to work, quality of life, and residual symptoms. Additionally, we compare these outcome parameters between NPSAH and PMSAH patients.

## Methods

### Patient population

Data of all consecutive SAH patients admitted to the department of Neurosurgery at the Amsterdam University Medical Centre, a tertiary referral center for the treatment of SAH patients in the Amsterdam metropolitan area with a total population of approximately 2.5 million people, are systematically recorded in a prospective SAH registry. For this study, we used data from all nSAH patients, admitted from January 2012 up to December 2023. SAH was diagnosed on a non-contrast CT-scan, magnetic resonance imaging (MRI) or by lumbar puncture (LP). Patients were excluded when a CT-angiography (CTA), Digital Subtraction Angiography (DSA), or Magnetic Resonance Angiography (MRA) was not obtained, or could not reliably be assessed. PMSAH was defined as a hemorrhage located in front of the brain stem, mainly in the interpeduncular cistern, with or without extension to the ambient, chiasmal, medial 1/3 part of the horizontal part of the Sylvian cisterns, on a CT-scan performed within 72 hours after onset ^24^. Patients with a PMSAH pattern underwent a CTA, however, a DSA was not routinely performed according to local guidelines. All other included nSAH patients were stratified as NPSAH. These patients received both a CTA and a DSA. Patients were excluded when a causative aneurysm for the hemorrhage, or other intracranial vascular pathology, was found. In DSA-negative patients, generally, a repeat DSA or MRI/MRA was performed approx. 14 days post-hemorrhage. Exceptions for a repeat DSA were made during interdisciplinary neurovascular boards, and included LP-positive SAH patients, and patients with severe atherosclerosis.

Following current (inter)national and local guidelines, nimodipine was administered in all NPSAH patients, but not in PMSAH patients. NPSAH patients were usually admitted at least until the repeat DSA was performed. After diagnosis at our center, PMSAH patients were only admitted to our center if the clinical status of a patient warranted observation in a neurosurgical treatment center. Routine clinical and radiological follow-up in SAH patients was done at six months after the hemorrhage.

### Data collection and definitions

Data collection was prospectively performed by a trained research nurse using a predefined Case Report Form (CRF). Castor Electronic Data Capture was used to capture collected data.

Demographic characteristics collected included: sex, age, history of smoking (yes/no), medical history, use of anticoagulant and/or antiplatelet medication (yes/no), World Federation of Neurosurgical Societies (WFNS) score at time of admission to the treatment center, and modified Fisher grade on the first non-contrast CT-scan. Relevant medical history involved intracranial hemorrhage (hemorrhagic cerebrovascular accidents, traumatic hemorrhages), hypertension, cardiovascular disease (ischemic cerebrovascular accident, cardiac events, and other vascular pathologies), diabetes mellitus, and diseases of the central nervous system.

Clinical course characteristics that were collected involved in-hospital mortality, and complications: recurrent bleeding, hydrocephalus (acute and chronic), delayed cerebral ischemia (DCI), seizures, infections (pneumonia, urine tract infection, and meningitis), and delirium. Definitions of the complications can be found in **supplement table 1** ^25–27^. All results were reported according to the STROBE guidelines.

### Outcome assessment

Outcome assessment is routinely performed by a trained research nurse, either during outpatient clinic appointments or through standardized telephone interviews with follow-up at six months after the hemorrhage. If six months follow-up was not available, we used the 12 months assessment if present. The research nurse is trained to assess the mRS. The other outcome measures involve questionnaires which are completed by patients themselves.

The mRS (score 0-6) is used to assess functional outcome regarding physical dependency and return to work ^28^. Quality of life is assessed using the EuroQol-5Dimension health questionnaire (EQ-5D) (maximum index-score 1,000) and EuroQol-visual analogue scale (EQ-VAS) (maximum score 100) and the hospital anxiety and depression scale (HADS) (maximum score per category anxiety 21/21 and depression 21/21) ^29–31^. For the EQ-5D and EQ-VAS, scores were compared to EQ-5D- and EQ-VAS reference scores for the Dutch population according to age ^29^. Over time, both EQ-5D-3L and EQ-5D-5L were used. Scores were pooled according to the ‘cross walk method’ ^30^. Furthermore, we assess for persistent symptoms such as tiredness, headache, forgetfulness, and anxiety following the hemorrhage by ‘yes or no’ institutional 14-item questionnaire.

### Statistical analysis

Neurological admission status was dichotomized in ‘good’ (WFNS-grade I-III) and ‘poor’ (WFNS-grade IV-V). Functional outcome was dichotomized in ‘favorable’ (mRS score 0-2) and ‘unfavorable’ (mRS score 3-6). Additionally, excellent functional outcome was defined as mRS-score 0-1. Return to work was dichotomized in ‘yes’ and ‘no’. Patients who were not employed at the time of the hemorrhage (unemployed, stay-at home, retired, or incapacitated) were excluded from the analysis. For HADS and EQ-5D/-VAS scores were reported. For the HADS, a score >7 for each subdomain was suggestive for (potential) anxiety or depression disorder ^31,32^.

Descriptive statistics were reported for all baseline characteristics, the clinical course characteristics, and outcomes at six-month follow-up for nSAH patients, and stratified per diagnosis (PMSAH and NPSAH). Continuous variables were assessed for normal distribution, with a Shapiro-Wilk value >0.9 defined as normally distributed. Normally distributed data were reported as means with standard deviations (SD) and as medians with interquartile ranges (IQR) for non-normally distributed data. Student’s t-test was done for normally distributed continuous variables and Mann-Whitney U test for non-normally distributed continuous variables, respectively. Dichotomous- and categorical variables, and outcome were compared using a Fisher’s exact test. Results with a *P* value <0.05 were considered statistically significant. We assessed the association between diagnosis and outcome and between diagnosis and return to work using a univariable logistic regression analysis. To detect confounders, a bivariable analysis was done with demographic variables added individually to the model. All demographic variables were considered potential confounding variables. Demographic variables that changed the crude odds ratio (OR) by more than 10%, were considered confounders. In case confounders were detected and a sufficient number of events in the smallest group of the dependent variable was observed, a multivariable model was built including the main outcomes as dependent variables (‘good outcome’ defined as mRS-score 0-2 and ‘return to work’) and confounders and diagnosis group as independent variables. All analyses were done using SPSS statistics, version 28.0.

## Results

### Patient characteristics and clinical course

A total of 325 nSAH patients (127 [39%] female; mean age 56 yrs.) were included in the present study, comprising 191 (59%) NPSAH and 134 (41%) PMSAH (**flowchart 1**). In DSA negative patients, a repeat DSA and/or MRI/MRA or CTA was performed in 115 (60%) NPSAH patients. Reasons for not performing a repeat DSA were: LP-positive SAH (n=39), the first DSA was performed several days (range 4 – 14 days) after the initial hemorrhage and was of sufficient quality (n=14), patient specific characteristics (e.g., severe atherosclerosis) (n=8), patients’ request (n=1), death prior to repeat imaging (n=5), and unknown reasons (n=9). One-hundred ninety-four (60%) nSAH patients were employed at the time of the hemorrhage (**table 1**). Acute hydrocephalus was the most commonly reported complication, affecting 71 patients (22%), with definitive shunting required in 11 patients (15%).

**Table 1.**
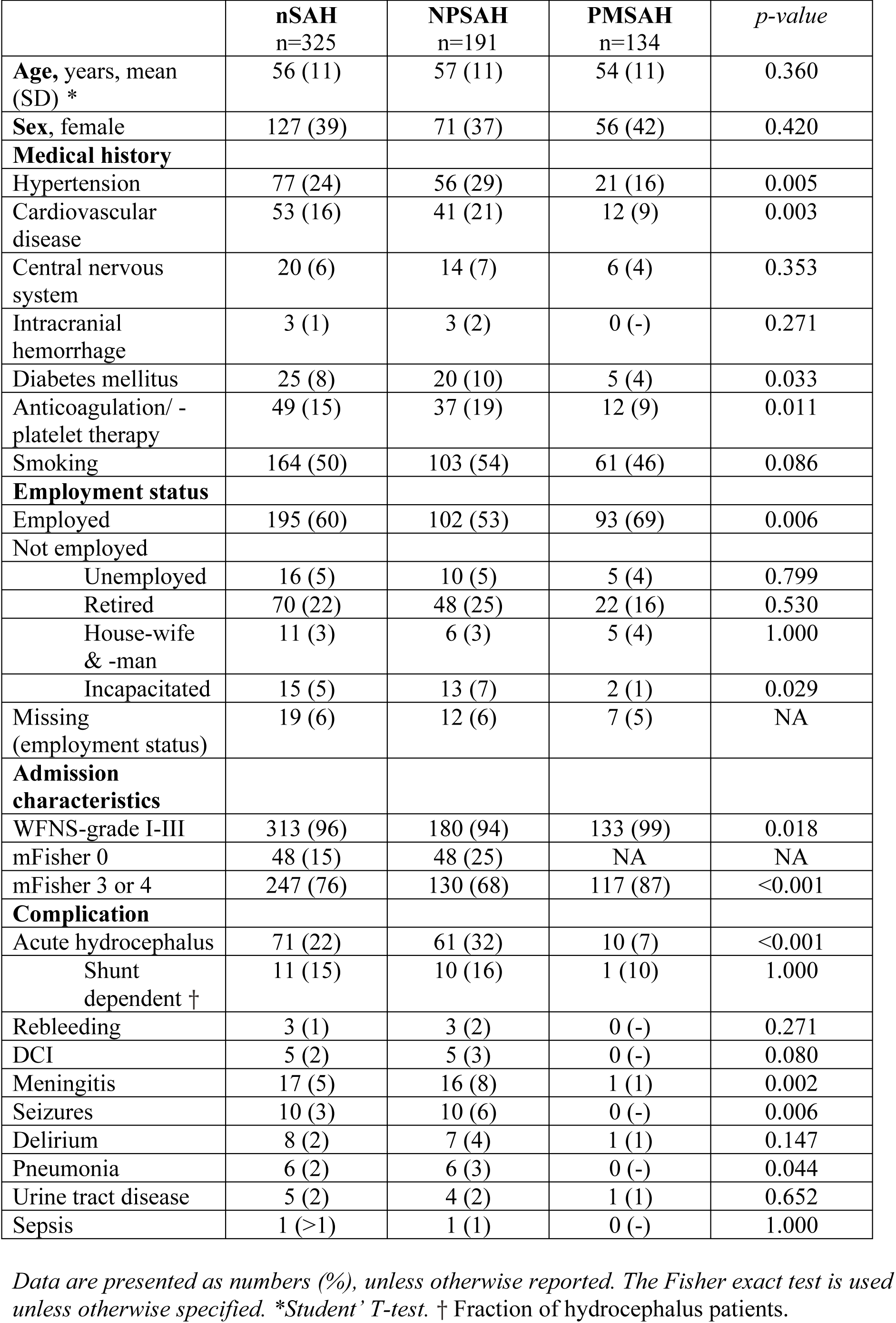
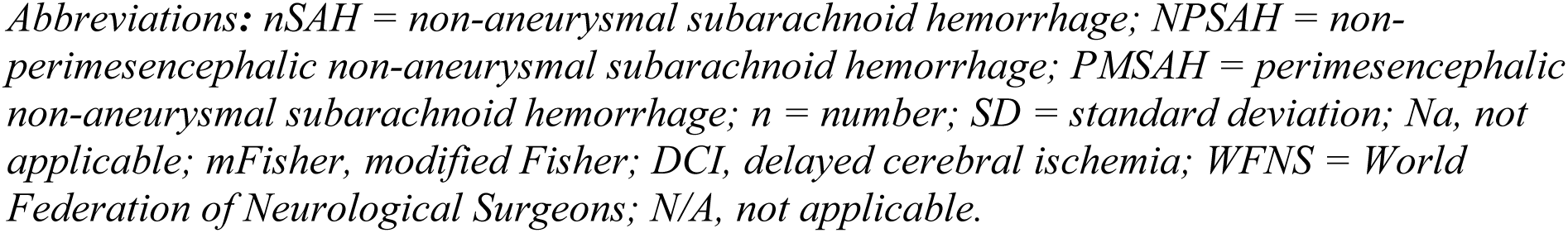
Baseline characteristics and clinical course of 325 nSAH patients.

|  | **nSAH**<br>n=325 | **NPSAH**<br>n=191 | **PMSAH**<br>n=134 | *p-value* |
| --- | --- | --- | --- | --- |
| **Age**, years, mean (SD) * | 56 (11) | 57 (11) | 54 (11) | 0.360 |
| **Sex**, female | 127 (39) | 71 (37) | 56 (42) | 0.420 |
| **Medical history** |  |  |  |  |
| Hypertension | 77 (24) | 56 (29) | 21 (16) | 0.005 |
| Cardiovascular disease | 53 (16) | 41 (21) | 12 (9) | 0.003 |
| Central nervous system | 20 (6) | 14 (7) | 6 (4) | 0.353 |
| Intracranial hemorrhage | 3 (1) | 3 (2) | 0 (-) | 0.271 |
| Diabetes mellitus | 25 (8) | 20 (10) | 5 (4) | 0.033 |
| Anticoagulation/ -platelet therapy | 49 (15) | 37 (19) | 12 (9) | 0.011 |
| Smoking | 164 (50) | 103 (54) | 61 (46) | 0.086 |
| **Employment status** |  |  |  |  |
| Employed | 195 (60) | 102 (53) | 93 (69) | 0.006 |
| Not employed |  |  |  |  |
| Unemployed | 16 (5) | 10 (5) | 5 (4) | 0.799 |
| Retired | 70 (22) | 48 (25) | 22 (16) | 0.530 |
| House-wife & -man | 11 (3) | 6 (3) | 5 (4) | 1.000 |
| Incapacitated | 15 (5) | 13 (7) | 2 (1) | 0.029 |
| Missing (employment status) | 19 (6) | 12 (6) | 7 (5) | NA |
| **Admission characteristics** |  |  |  |  |
| WFNS-grade I-III | 313 (96) | 180 (94) | 133 (99) | 0.018 |
| mFisher 0 | 48 (15) | 48 (25) | NA | NA |
| mFisher 3 or 4 | 247 (76) | 130 (68) | 117 (87) | <0.001 |
| **Complication** |  |  |  |  |
| Acute hydrocephalus | 71 (22) | 61 (32) | 10 (7) | <0.001 |
| Shunt dependent † | 11 (15) | 10 (16) | 1 (10) | 1.000 |
| Rebleeding | 3 (1) | 3 (2) | 0 (-) | 0.271 |
| DCI | 5 (2) | 5 (3) | 0 (-) | 0.080 |
| Meningitis | 17 (5) | 16 (8) | 1 (1) | 0.002 |
| Seizures | 10 (3) | 10 (6) | 0 (-) | 0.006 |
| Delirium | 8 (2) | 7 (4) | 1 (1) | 0.147 |
| Pneumonia | 6 (2) | 6 (3) | 0 (-) | 0.044 |
| Urine tract disease | 5 (2) | 4 (2) | 1 (1) | 0.652 |
| Sepsis | 1 (>1) | 1 (1) | 0 (-) | 1.000 |

Data are presented as numbers (%), unless otherwise reported. The Fisher exact test is used unless otherwise specified. \*Student' T-test. † Fraction of hydrocephalus patients.
*Abbreviations: nSAH = non-aneurysmal subarachnoid hemorrhage; NPSAH = non-perimesencephalic non-aneurysmal subarachnoid hemorrhage; PMSAH = perimesencephalic non-aneurysmal subarachnoid hemorrhage; n = number; SD = standard deviation; Na, not applicable; mFisher, modified Fisher; DCI, delayed cerebral ischemia; WFNS = World Federation of Neurological Surgeons; N/A, not applicable.*

### Outcome nSAH

Follow-up data were available for 303 patients (93%), comprising 179 (94%) and 124 (93%) patients of the NPSAH and PMSAH population, respectively (**table 2**). Reasons for missing follow-up at six months were relocation to another region or country (n=5), at patients’ request (n=5), and unknown (n=12). Seven (2%) patients died between hemorrhage and follow-up, all during hospital stay. The reported causes of death included refraining from further treatment in cases of persisting coma due to the hemorrhage (n=4), refraining from further treatment in an elderly patient with hydrocephalus (n=1), due to respiratory failure (n=1), and multi-organ failure (n=1). In 11 patients (4%), a 12 months follow-up was used due to absence of a six months assessment.

**Table 2.** Outcome at six months’ follow-up in 303 nSAH patients.

|  | **nSAH** | **NPSAH** | **PMSAH** | *p-value* |
| --- | --- | --- | --- | --- |
| **Follow-up** | 303 (93) | 179 (94) | 124 (93) | 0.662 |
| Loss to follow/up | 22 (7) | 12 (6) | 10 (7) | NA |
| **Follow-up time**, median (Q1/Q3), months | 6 (5.9/6.4) | 5.9 (5.8/6.3) | 6.2 (6.0/6.5) |  |
| **Functional outcome**, mRS |  |  |  |  |
| All-cause mortality at 6 months (mRS 6) | 7 (2) | 6 (3) | 1 (1) | 0.246 |
| Favorable (mRS-score 0-2) | 271 (89) | 157 (88) | 114 (92) | 0.260 |
| Excellent (mRS-score 0-1) | 120 (40) | 63 (35) | 57 (46) | 0.073 |
| **Functional outcome**, return to work * |  |  |  |  |
| Returned to work | 141 (77) | 67 (70) | 74 (85) | 0.001 |
| a. Fully returned to work | 71 (39) | 30 (31) | 41 (47) | 0.034 |
| b. Partially returned to work | 70 (38) | 37 (39) | 33 (38) | 1.000 |
| Not returned to work | 42 (23) | 28 (29) | 14 (15) | 0.026 |
| **Quality of life** |  |  |  |  |
| EQ-5D, mean score (SD) † | 0.827 (0.184) | 0.825 (0.183) | 0.830 (0.187) | 0.848 |
| EQ-VAS, mean score (SD) ‡ | 74 (16) | 74 (17) | 75 (15) | 0.363 |
| HADS total, mean score (SD) § | 9.4 (8) | 9.5 (8) | 8.9 (8) | 0.405 |
| Anxiety, mean score (SD) | 5.2 (4) | 4.9 (4) | 5.5 (4) | 0.317 |
| Anxiety deviant (score >7) | 53 (23) | 35 (15) | 18 (8) | 0.340 |
| Depression, mean score (SD) | 4.2 (4.2) | 4.3 (4) | 4.0 (4) | 0.582 |
| Depression deviant (score >7) | 48 (21) | 32 (14) | 16 (7) | 0.323 |
| **Residual symptoms** \|\| |  |  |  |  |
| One or more residual symptoms | 202 (84) | 125 (86) | 77 (81) | 0.284 |

*Data are presented as numbers (%), unless otherwise reported. Abbreviations: SD, standard deviation; IQR, interquartile range; mRS, modified Rankin Scale; HADS, hospital anxiety and depression scale; EQ-5D, EuroQol-5 Dimensions. The Fisher exact test is used unless otherwise specified. \*Calculated over 183 (96 NPSAH and 87 PMSAH) patients of which the employment status was known after the hemorrhage. †Calculated over 240 (141 NPSAH and 99 PMSAH) patients. ‡Calculated over 231 (139 NPSAH and 92 PMSAH) patients. § Calculated over 235 (141 NPSAH and 94 PMSAH) patients. || Calculated over 241 (146 NPSAH and 95 PMSAH) patients.*

A favorable functional outcome (mRS-score 0-2) was reported in 271/303 patients (89%) at a median (IQR) follow-up duration of six (5.9 - 6.4) months (**figure 2**). Excellent outcome (mRS-score 0 – 1) was reported in 120 patients (40%). At six months follow-up, 141/183 (77%) patients returned to work (return to work was missing in 12 patients who were lost to follow-up) (**figure 3**). Fully returning to the previous level (same position and workload) of work was reported in 71/183 patients (39%).

**Figure 1.**
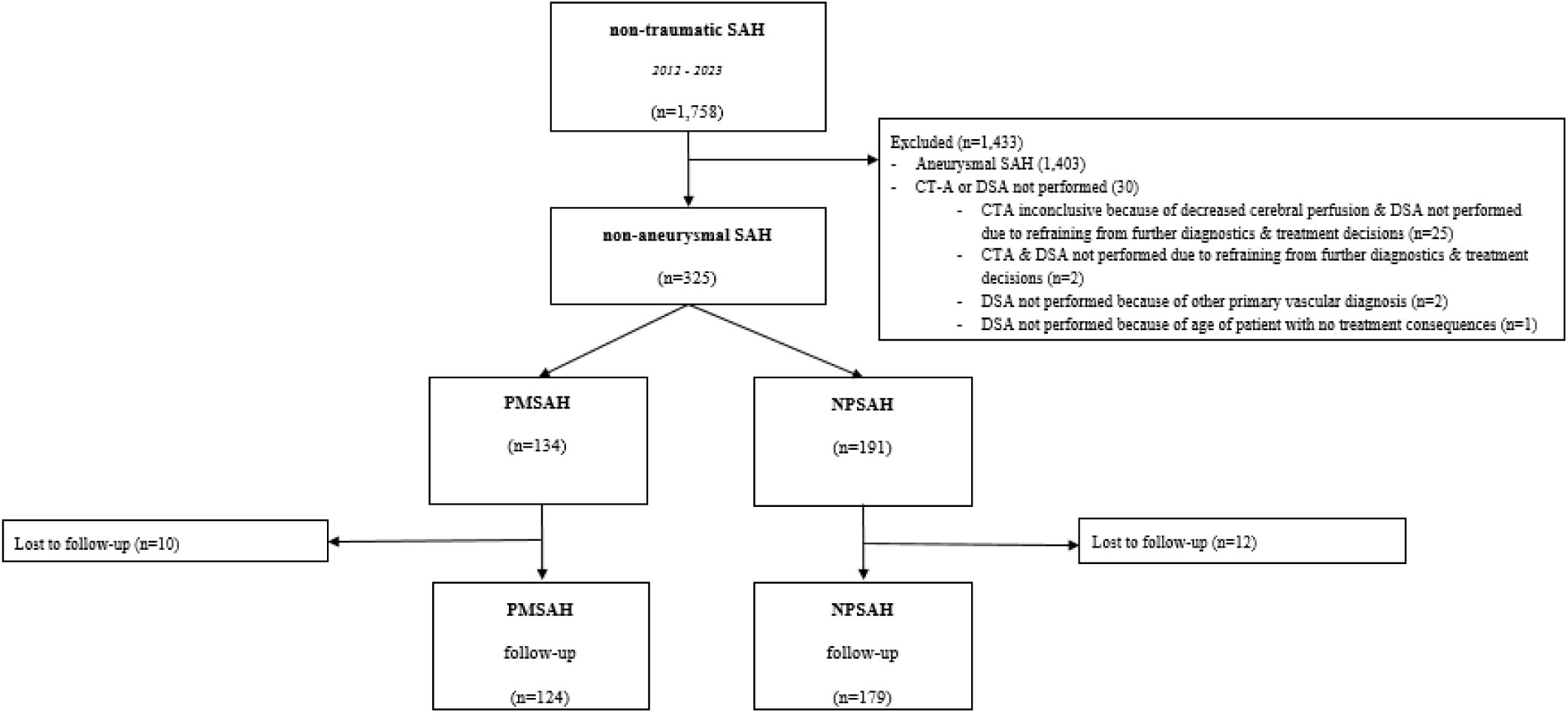
Flowchart of included and excluded patients.

**Figure 2.**
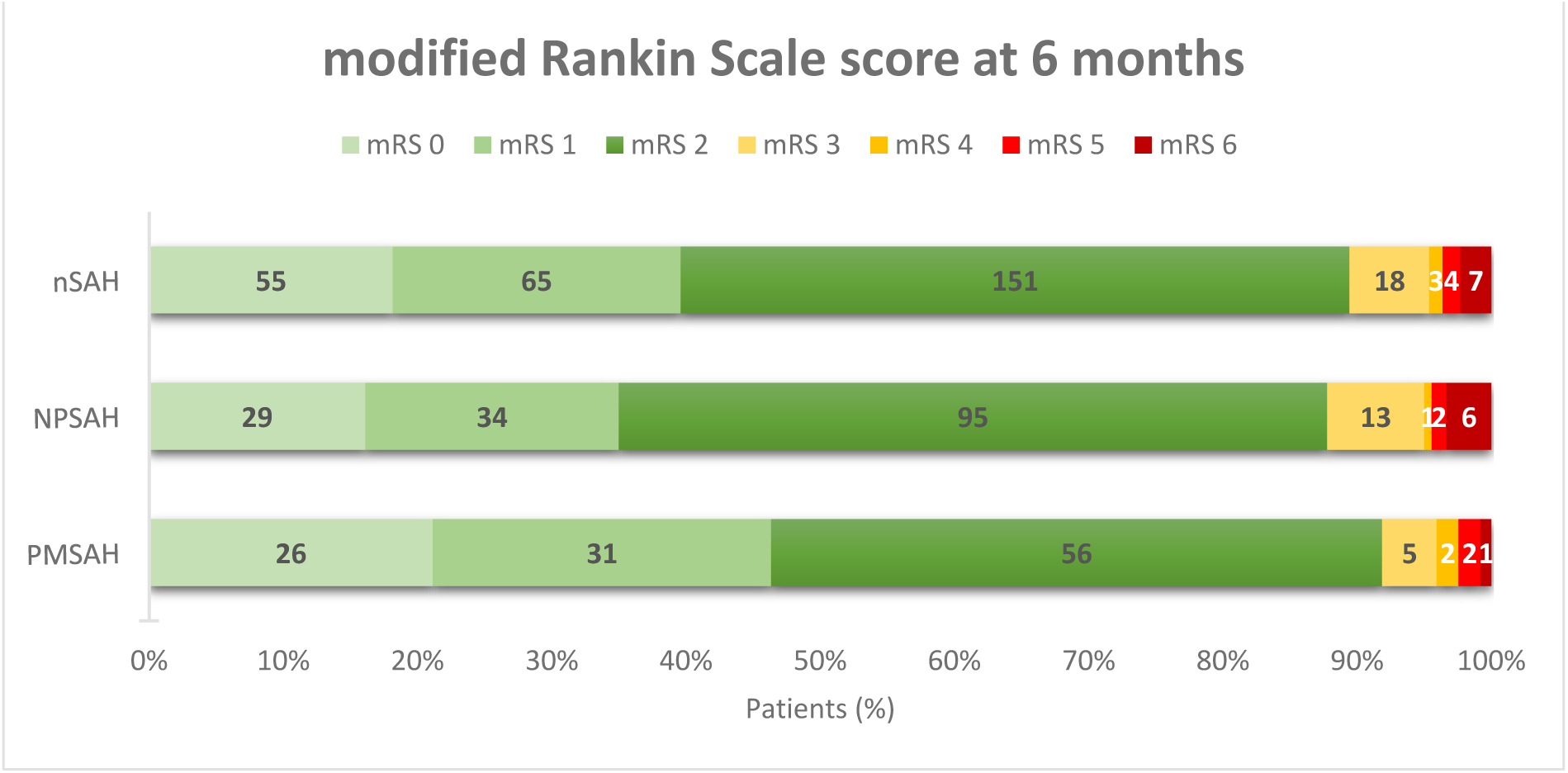
Functional outcome at six months follow-up.

**Figure 3.**
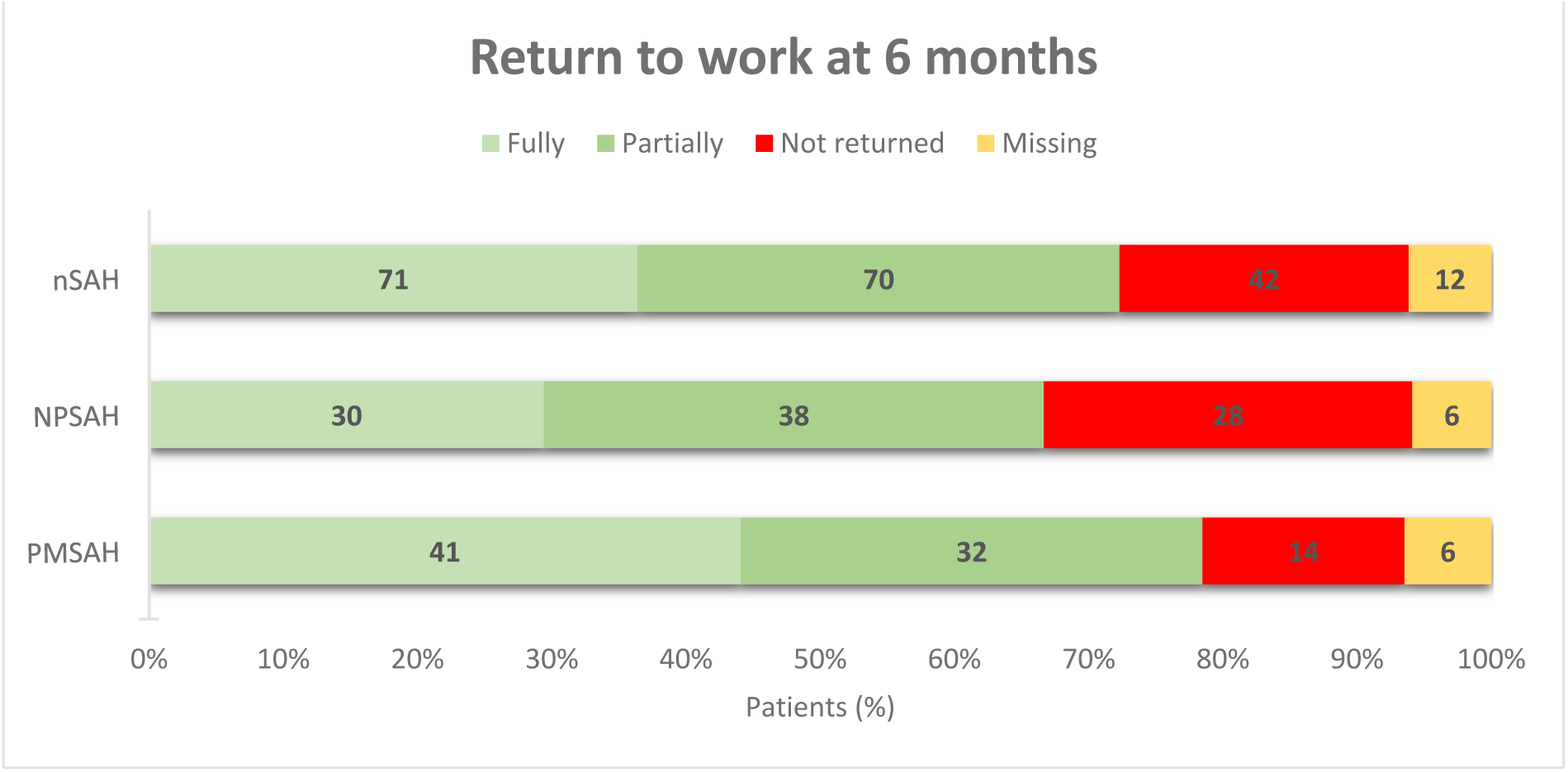
Return to work at six months follow-up.

Symptoms of increased anxiety (HADS-A) and depression (HADS-D) were present in 53/234 (23%) and 49/235 (21%) patients. In 38/234 patients (16%), both HADS-A & -D were increased. Residual symptoms, based on the institutional PROMS 14-item questionnaire, were present in 202/241 patients (84%). Only 39 patients (16%) denied experiencing residual symptoms. The most frequently reported symptoms included earlier fatigue (n=164; 68%), increased difficulties in concentration (n=130; 54%), and increased forgetfulness (n=121; 50%) (**supplement table 2**).

### NPSAH vs. PMSAH

NPSAH patients were older, suffered from various comorbidities and were less often employed at the time of the hemorrhage (**table 1**). Acute hydrocephalus was reported in 61 NPSAH patients (32%) and 10 PMSAH patients (7%) (P<0.001). NPSAH patients were more likely to develop a meningitis (p=.002). Rebleeding and delayed cerebral ischemia exclusively occurred in NPSAH patients. Favorable functional outcome (mRS-score 0-2) was achieved in 157 NPSAH (88%) and 114 PMSAH (92%) patients (p=.260) (**table 2**). PMSAH patients were more likely to return to work at six months (68 [71%] NPSAH and 73 [84%] PMSAH, respectively, p<.01). Furthermore, PMSAH patients were more likely to fully return to work (30 [31%] NPSAH, 41[47%] PMSAH, respectively, p=.034). No differences were found in EQ-5D, EQ-VAS, and HADS scores. NPSAH patients were more prone for developing increased fatigue (p=.0048) (**supplement table 2**). Subgroup analysis leaving out the 48 NPSAH mFisher 0 (LP-positive SAH) showed no differences in functional outcome or quality of life.

### Multivariable analysis

Confounding effects were analyzed for all demographic variables. For good outcome, (mRS-score 0-2), age, hypertension, cardiovascular disease, DM2, and the use of anticoagulant or – platelet therapy were considered confounders. However, a multivariable analysis could not be performed due to the insufficient number of events in the smallest group of the dependent variable (patients with an unfavorable outcome), which precluded the construction of a stable multivariable model. No confounders were identified for returning to work.

## Discussion

This large prospective, single-center, observational study of non-aneurysmal, non-traumatic subarachnoid hemorrhage patients reveals that the majority of patients did not return to their previous level of work despite that most patients reported a good outcome (mRS 0-2) at six months follow-up. Furthermore, this study shows that residual symptoms, such as increased fatigue and concentration difficulties, are commonly reported. These findings may nuance the perception of what has been previously been supposed as good outcome in these patients.

Outcome in patients after SAH has been extensively studied, often focusing on physical dependency ^5,11,13^. The assessment of functional outcome in the present study was done using the mRS. Alike previous studies, the majority of patients achieved a favorable (mRS-score 0-2) outcome ^6^. The majority of nSAH patients in this study had an mRS-score 2, reflecting slight disability but not returning to all previous daily activities. The standardized mRS telephone interview assesses whether patients were employed at the time of the hemorrhage, and whether they experience problems with their work at follow-up. Patients who do not return to their previous level of work or do not return to work at all are reported as an mRS-score 2. In the present study, the majority of the patients who were employed at the time of the hemorrhage did not fully return to work, or did not (yet) return to work at all at six months follow-up. This prompts the question of whether resuming prior daily activities, including returning to work, can be used as a criterion for favorable outcome, and how an ‘excellent outcome’ might be defined. In previous studies, the outcome for nSAH patients may have been considered excellent by comparing it to aSAH in terms of physical dependency and mortality ^6,9^. However, studies comparing aSAH and nSAH cohorts remain sparse ^3,33–35^. For aSAH, an mRS-score 0-2 or even 0-3 is a commonly used, an broadly accepted cut-off for defining favorable outcome ^9,36^. For nSAH, a favorable (functional) outcome may better be defined as mRS-score 0-1, as argued by Patel et al. ^37^. In the present study, about 40% of nSAH patients reported an mRS-score 0-1. From this perspective, fewer than half of the patients report an ‘excellent outcome’ at six months follow-up. Furthermore, the majority of the patients report one or more residual symptoms, which implicates that outcome involves more than (physical) dependency. Presence of these symptoms at follow-up may to some extent explain why patients did not (fully) return to their work or resuming former daily activities, and the proportion of patients reporting a decreased quality of life, or symptoms of anxiety and/or depression. These findings nuance the relatively benign clinical course of nSAH patients.

As a personalized post-hemorrhage support system is becoming increasingly important, the findings of the present study could contribute for the counselling of these patients. In this regard, early recognition of deficits during follow-up in patients may increase the attention for implementing additional interventions. For the follow-up of SAH patients at our center, we use the 14-item questionnaire to address various symptoms that have been previously mentioned by patients underlining the importance of using patient-reported outcome measures (PROMS). The use of PROMS has been previously studied in stroke ^38–42^. It was found that although patients may report a favorable functional outcome on the mRS-score, patients could still report other complaints resulting in an overall unfavorable outcome. ^38,42^ By using PROMS as a screening tool during follow-up, nSAH patients that may require additional follow-up or care after discharge may be more easily recognized and specific interventions could be initiated accordingly. One such intervention could be rehabilitation. Previous studies have already addressed the possible positive influence of rehabilitation on recovery in nSAH patients ^43,44^. Findings of the present study regarding not (fully) returning to work and presence of residual symptoms underline the possible relevance of rehabilitation in these patients and warrants further investigation in larger prospective studies.

The present study holds some strengths and limitations. Strengths involve the prospective nature of the study including a structured follow-up assessment by a trained research nurse as well as the large sample size compared to previous (prospective) studies ^8,13–15,18–23^. Furthermore, less than 10% of all patients were lost to follow-up. In the present study, we used validated (mRS, EQ-5D, EQ-VAS, and HADS) and non-validated PROMS (14-item questionnaire) tools for assessing outcome in multiple dimensions. A limitation involved the number of missing data for the HADS, EQ-5D, and the 14-item residual symptoms questionnaire. A possible explanation is that follow-up, especially since the COVID-19 pandemic, was increasingly done through telephone instead of in-person consults. This could have made assessment of the questionnaires more challenging and therefore lead to a higher likelihood of missing data and introduced bias in the reported findings. Selection bias may be present if patients are physically not able to fill out questionnaires, resulting in an overestimation of good outcome in the study population. Another limitation is that PMSAH patients were not routinely admitted to our center after diagnosis and adverse events at the referring hospital may not have been communicated to our center. However, since our center serves as the main tertiary referral center for SAH care, chances of adverse events not being communicated to our center are expected to be small. The lack of information regarding the reason of not (fully) returning to work can be considered another limitation. It remains unclear whether not fully returning to work was due the physical and/or psychological impairments after the hemorrhage or whether it was the patients’ own decision to ease off after the hemorrhage and to retire (earlier).

## Conclusion

The majority of patients did not yet (fully) return to their previous level of work, despite reporting a favorable mRS 0-2 score, at six months follow-up after non-aneurysmal, non-traumatic subarachnoid hemorrhage. Moreover, most patients report long-lasting residual symptoms. These findings nuance the proposed ‘good outcome’ in non-aneurysmal subarachnoid hemorrhage patients and warrants further research. This could focus on prospectively assessing outcome using PROMS in larger sample sizes at long-term follow-up as well as studying the influence of rehabilitation on recovery and returning to work.

## Acknowledgments

Not applicable.

## Data Availability Statement

All data requests should be submitted to Dagmar Verbaan for consideration. Access to anonymised data may be granted following review.

## Miscellaneous Disclosures

This study was waived for ethical approval by the Institutional Review Board of the Amsterdam UMC because it did not fall under the scope of the Medical Research Involving Human Subjects Act (WMO). This study was completed in accordance with the Helsinki Declaration as revised in 2013.

## Supplement tables & figures

**Supplement table 1.** Definitions of various clinical terms.

| **Clinical term** | **Definition** |
| --- | --- |
| **Recurrent bleeding**<br>(rebleeding) | A second (or third etc.) bleeding after the initial bleeding. Rebleeding was either diagnosed with a non-contrast CT-scan or in case of high clinical suspicion (e.g., acute clinical deterioration combined with abrupt increase in blood pressure, bradycardia, respiratory alterations, or the appearance of a sudden increase in production of CSF with fresh blood through ventricular drainage. |
| **Acute hydrocephalus** | Enlarged ventricles on imaging, assessed by an experienced neuro-radiologist, or by increased intracranial pressure diagnosed by lumbar puncture or ventricular catheter placement or by increased intracranial pressure diagnosed through a lumbar puncture or ventricular catheter placement. |
| **Chronic hydrocephalus** | Requiring definitive CSF-shunting. |
| **Delayed cerebral ischemia (DCI)** | The occurrence of focal neurological impairment (such as hemiparesis, aphasia, apraxia, hemianopia, or neglect), or a decreased of at least 2 points on the Glasgow Coma Scale. This should last for at least 1 hour, is not apparent immediately after aneurysm occlusion, and cannot be attributed to other causes by means of clinical assessment, CT or magnetic imaging scanning of the brain, and appropriate laboratory studies. |
| **Seizures** | Clinical appearance of rhythmic tonic or clonic movements for which anti-epileptic drugs were started or by electroencephalography (EEG). |
| **Pneumonia** | Clinical symptoms followed by a positive culture the microbiological analysis of sputum or a consolidation on the chest X-ray or when antibiotics were prescribed specifically for this indication. |
| **Urinary tract infection** | Clinical symptoms followed by a positive culture based on the microbiological analysis of urine or leukocytosis or bacteremia in the urine sediment. |
| **Meningitis** | Clinical symptoms followed by a positive culture based on the microbiological analysis of CSF or when antibiotics were prescribed specifically for this indication. |
| **Delirium** | In case of treatment with haloperidol and patients had a delirium observation screening (DOS) score >4 or if the Richmond Agitation Sedation Scale (RASS) was scored between +4 and -4. |

**Supplement table 2.** 14-item questionnaire.

| **Questionnaire item** | **nSAH** | **NPSAH** | **PMSAH** | *p-value* |
| --- | --- | --- | --- | --- |
| Q1. Increased fatigue * | 164 (68) | 106 (73) | 58 (60) | 0.048 |
| Q2. More sensitive for loud noises * | 111 (46) | 62 (43) | 49 (51) | 0.236 |
| Q3. Increased forgetfulness * | 121 (50) | 78 (54) | 43 (45) | 0.189 |
| Q4. Increased concentration difficulties * | 130 (54) | 77 (53) | 53 (55) | 0.792 |
| Q5. Increased headache * | 100 (42) | 62 (43) | 38 (40) | 0.689 |
| Q6. Increased insomnia † | 50 (21) | 30 (21) | 20 (21) | 1,000 |
| Q7. Increased daytime sleepiness * | 112 (47) | 74 (51) | 38 (40) | 0.088 |
| Q8. Worse or blurred vision ‡ | 66 (28) | 37 (26) | 29 (30) | 0.465 |
| Q9. Increased dizziness * | 67 (28) | 42 (29) | 25 (26) | 0.661 |
| Q10. Increased excitable § | 92 (38) | 54 (38) | 38 (40) | 0.787 |
| Q11. Increased anxious feelings * | 84 (35) | 54 (37) | 30 (31) | 0.408 |
| Q12. Increased gloominess * | 60 (25) | 39 (27) | 21 (22) | 0.447 |
| Q13. Increased balance problems * | 57 (24) | 36 (25) | 21 (22) | 0.644 |
| Q14. Increased lack of initiative \|\| | 67 (28) | 42 (30) | 25 (26) | 0.660 |

\* Calculated over 241 (145 NPSAH and 96 PMSAH) patients. † Calculated over 239 (144 NPSAH and 95 PMSAH) patients. ‡ Calculated over 239 (143 NPSAH and 96 PMSAH) patients. § Calculated over 240 (144 NPSAH and 96 PMSAH) patients. || Calculated over 238 (142 NPSAH and 96 PMSAH) patients.

